# Decreased tissue stiffness in glioblastoma by MR Elastography is associated with increased cerebral blood flow

**DOI:** 10.1101/2021.06.11.21258742

**Authors:** Siri Fløgstad Svensson, Elies Fuster-Garcia, Anna Latysheva, Jorunn Fraser-Green, Wibeke Nordhøy, Omar Isam Darwish, Ivar Thokle Hovden, Sverre Holm, Einar O. Vik-Mo, Ralph Sinkus, Kyrre Eeg Emblem

## Abstract

**PURPOSE:** Understanding how mechanical properties relate to functional changes in glioblastomas may help explain different treatment response between patients. The aim of this study was to map differences in biomechanical and functional properties between tumor and healthy tissue, to assess any relationship between them and to study their spatial distribution.

**METHODS:** Ten patients with glioblastoma and 17 healthy subjects were scanned using MR Elastography, perfusion and diffusion MRI. Stiffness and viscosity measurements G′ and G′′, cerebral blood flow (CBF), apparent diffusion coefficient (ADC) and fractional anisotropy (FA) were measured in patients’ contrast-enhancing tumor, necrosis, edema, and gray and white matter, and in gray and white matter for healthy subjects. A regression analysis was used to predict CBF as a function of ADC, FA, G′ and G′′.

**RESULTS:** Median G′ and G′′ in contrast-enhancing tumor were 13% and 37% lower than in normal-appearing white matter (P<0.01), and 8% and 6% lower in necrosis than in contrast-enhancing tumor, respectively (P<0.05). Tumors showed both inter-patient and intra-patient heterogeneity. Measurements approached values in normal-appearing tissue when moving outward from the tumor core, but abnormal tissue properties were still present in regions of normal-appearing tissue. Using both a linear and a random-forest model, prediction of CBF was improved by adding MRE measurements to the model (P<0.01).

**CONCLUSIONS:** The inclusion of MRE measurements in statistical models helped predict perfusion, with stiffer tissue associated with lower perfusion values.

## Introduction

Glioblastoma (GBM) is the most common primary malignant tumor in the central nervous system, and is usually rapidly fatal [1]. With a standard treatment regimen of surgery and radiochemotherapy the median survival time is only 12-15 months [1].

Perfusion and diffusion magnetic resonance imaging (MRI) are considered important methods for understanding the tumor biology and quantifying physiological processes [2]. However, the changing biomechanical properties of a solid tumor and its surrounding tissue *in vivo* are less studied. Moreover, understanding how the mechanical properties of tissue relate to functional changes, may help explain the substantial variation in appearance and treatment response between patients.

MR Elastography (MRE) is a technique for non-invasively measuring the biomechanical properties of tissue [3]. Earlier studies have found that glioma tumors differ from the healthy brain in terms of stiffness and viscosity [4-7]. Most studies on MRE in brain tumors present mean values for the tumor stiffness, yet glioblastomas display a large degree of heterogeneity, both between patients, and within the tumor [8]. Furthermore, glioblastomas are known to infiltrate the surrounding tissue beyond the contrast-enhancing tumor, and this peritumoral area appears to play a key role in tumor growing and recurrence [9].

In this study, we combine MRE of GBM patients with diffusion and perfusion imaging. We have segmented tumors into contrast-enhancing and necrotic regions, and FLAIR-enhanced edema region in order to study tumor heterogeneity and infiltration. The objective of our study is to map the differences between tumor and healthy tissue with regard to biomechanical and functional properties, to study their spatial distribution, and to assess any possible relationship between biomechanical and functional parameters.

## Materials and methods

### Acquisition

Data was collected from 10 patients, 5 females and 5 males (44-74 years, median 55 years), prior to any treatment. All tumors were IDH-wildtype glioblastomas. In addition, 17 healthy subjects were scanned, 8 females and 9 males (21-34 years, median 25 years). All subjects signed an informed consent form. The study was approved by the Norwegiean National Research Ethics Committee and the Oslo University Hospital review board. The examination was tolerated well by all subjects. Apart from the subjects included in the study, three healthy subjects and two patients were excluded due to inadequate MRE data quality.

The scans were performed on a 3T clinical MRI scanner (Ingenia, Philips Medical Systems, Best, the Netherlands) using a 32-channel head coil. Scan parameters for all sequences used are presented in **Table 1**. The MRE was performed using a gravitational transducer [10] attached on the side of the subject’s head to induce shear waves of 50 Hz into the brain. Further details about the acquisition can be found in [11].

**Table 1:**
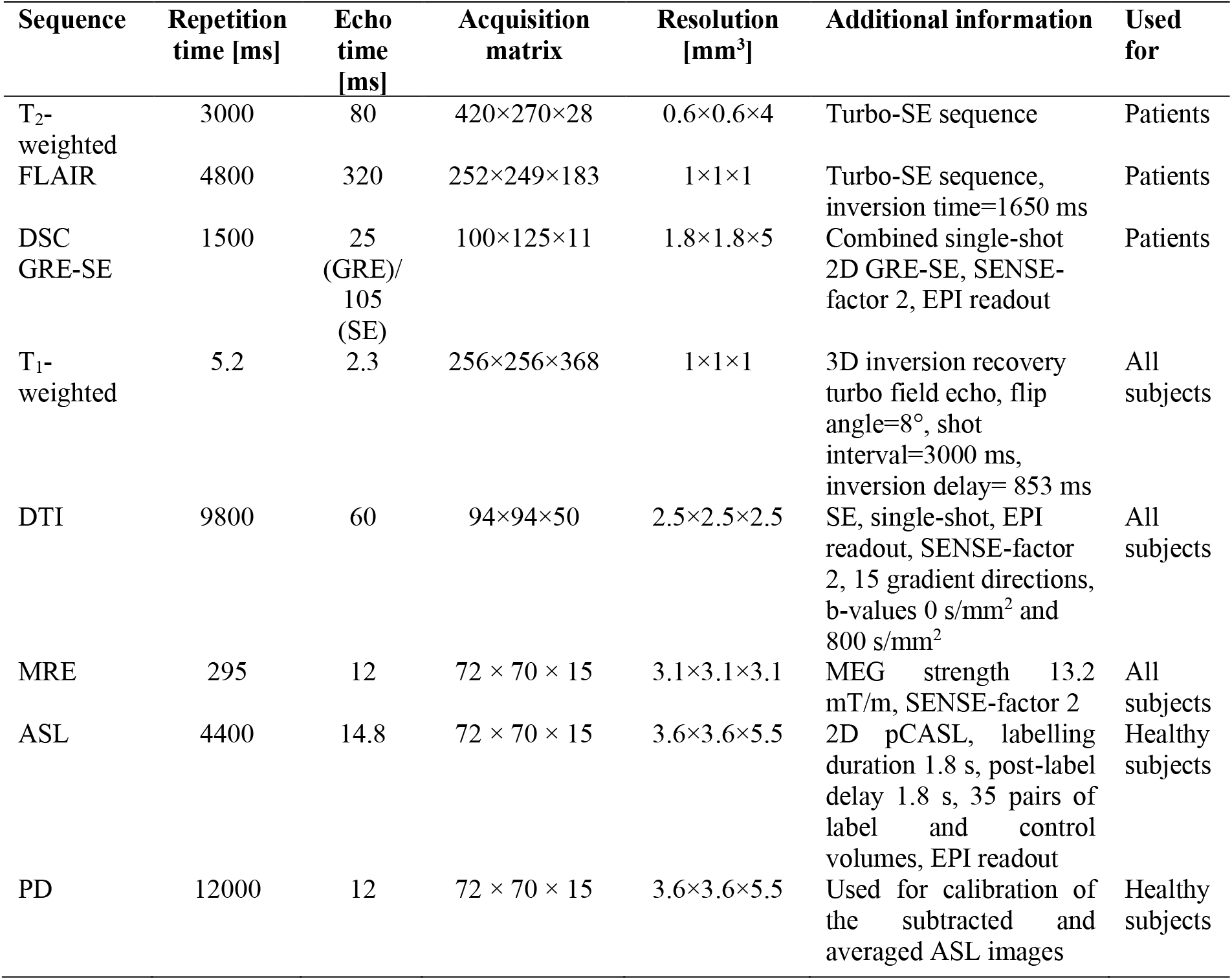
Scan parameters for patients and healthy subjects. Abbreviations: Gradient-echo (GRE), spin-echo (SE), echo-planar imaging (EPI), sensitivity encoding (SENSE), Diffusion tensor imaging (DTI), pseudo-Continuous Arterial Spin Labelling (pCASL), Proton-density weighted (PD).

### Image processing

Figure 1 shows resulting maps of cerebral blood flow (CBF), apparent diffusion coefficient (ADC) and fractional anisotropy index (FA) for both patients and healthy subjects. MRE produced maps of the shear storage modulus G′ (as a measure for stiffness) and loss modulus G′′ (related to the viscosity, meaning the tissue’s ability to dissipate energy) (**Fig. 1E-F, L-M**).

Patient data was segmented into contrast-enhancing tumor, edema, necrosis, and normal-appearing gray and white matter (**Fig. 1N**). Data from healthy subjects was segmented into gray and white matter. Gray matter was further subdivided into the deep gray and cortical regions.

**Figure 1:**
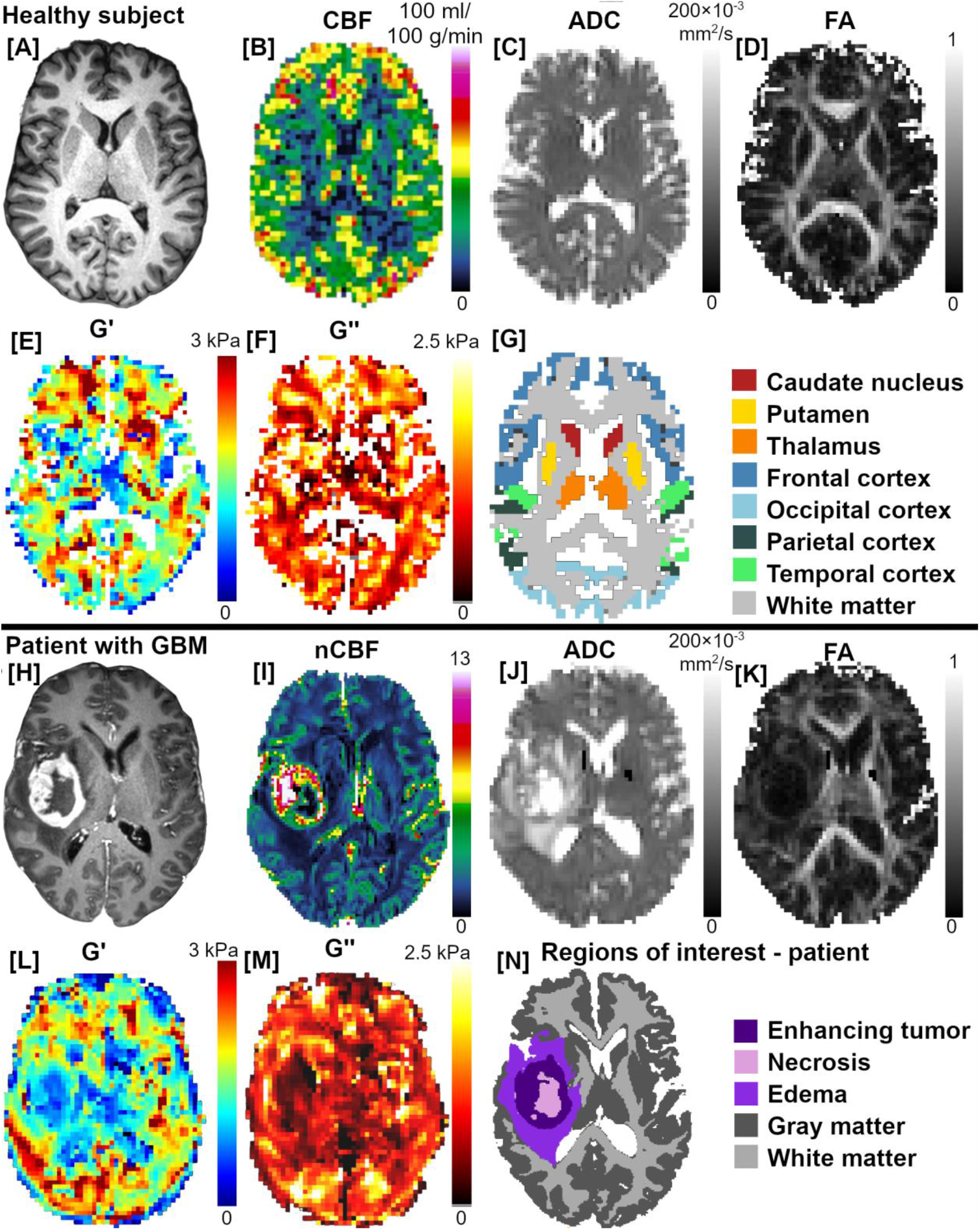
Image example of a healthy subject and patient. A) T1-weighted image, B) CBF, C) ADC, D) FA E) G′, and F) G′′, maps for a healthy subject. G) ROIs used for the healthy subjects, except from hippocampus (not visible in this slice). H) Contrast-enhanced T1-weighted image, I) normalized CBF, J) ADC, K) FA, L) G′, M) G′′ maps for a patient with glioblastoma. N) Patient ROIs.

Deep gray matter regions included in the study were head of the caudate nucleus, putamen, thalamus and hippocampus. Cortical gray matter regions included were the frontal, occipital, parietal and temporal lobe (**Fig. 1G**). In addition to absolute measurements, elastography and diffusion measurements in gray matter regions were normalized to the mean value in each subject’s white matter. Details about image processing and segmentation can be found in Supplementary information.

### Statistics

For each individual, we computed the mean value of each measurement in all regions of interest (ROIs). In the results below, we present the median of these mean values across individuals, with the range in parenthesis. Measurements in different regions were compared using a Wilcoxon signed-rank test. The relationship between the different parameters for mean values in each ROI was assessed by a Spearman rank-order test.

To visualize each tissue’s signature across parameters, mean measurements in each region for each subject was dimensionally reduced using t-distributed stochastic neighbor embedding (t-SNE).

Voxel-wise regression analysis was performed using both a simple linear model and a random forest model for perfusion as a function of ADC, FA, G′ and G′′. The performance of the regression models was evaluated by their root-mean-square error (RMSE) with a leave-one-patient-out cross-validation strategy. The different models were compared using a Wilcoxon signed-rank test.

A significance level of P<0.05 was assumed for all tests, after Holm-Bonferroni corrections for multiple comparisons. All statistical analyses were done using Matlab (version R2021a, MathWorks, Natick, MA, USA).

## Results

### Glioblastoma tissue is structurally degraded compared to healthy tissue

The results of the measurements in the different ROIs are summarized in **Table 2**.

**Table 2:** Measurements in patients (n=10) and healthy subjects (n=17). Upper part: G′, G′′, G′_norm_, G′′_norm (_normalized to each subject’s cNAWM), ADC, FA, and normalized CBF for patients. Lower part: Corresponding measurements in healthy subjects’ ROIs. CBF measurements in healthy subjects were acquired using ASL and are not directly comparable to patient CBF (using DSC).

| PATIENTS | Median<br>G' [kPa]<br>(range) | Median<br>G'' [kPa]<br>(range) | Median<br>G' norm<br>(range) | Median<br>G'' norm<br>(range) | Median<br>ADC<br>[10<sup>-3</sup> mm<sup>2</sup>/s]<br>(range) | Median<br>FA<br>(range) | Median<br>nCBF<br>(range) |
| --- | --- | --- | --- | --- | --- | --- | --- |
| Contrast-enhancing tumor | 1.40<br>(1.15-1.62) | 0.66<br>(0.55-0.80) | 0.86<br>(0.70-1.02) | 0.66<br>(0.51-0.83) | 1.21<br>(1.00-1.47) | 0.19<br>(0.14-0.25) | 3.77<br>(2.98-9.64) |
| Necrotic region | 1.15<br>(0.70-1.76) | 0.63<br>(0.37-0.81) | 0.70<br>(0.50-1.11) | 0.63<br>(0.40-0.74) | 1.36<br>(0.94-1.90) | 0.10<br>(0.09-0.16) | 2.14<br>(1.48-3.61) |
| Edema | 1.61<br>(1.43-2.16) | 0.88<br>(0.68-1.09) | 1.01<br>(0.86-1.33) | 0.90<br>(0.66-1.03) | 1.25<br>(1.01-1.47) | 0.20<br>(0.16-0.34) | 1.50<br>(0.97-2.46) |
| Average gray matter | 1.65<br>(1.34-1.73) | 1.02<br>(0.90-1.16) | 1.01<br>(0.87-1.07) | 0.99<br>(0.91-1.09) | 0.91<br>(0.85-1.08) | 0.16<br>(0.14-0.17) | 2.39<br>(2.06-2.94) |
| Average white matter | 1.61<br>(1.36-1.90) | 1.04<br>(0.91-1.23) | - | - | 0.82<br>(0.77-0.89) | 0.39<br>(0.34-0.43) | 1.29<br>(1.11-1.45) |
| HEALTHY SUBJECTS | Median<br>G' [kPa]<br>(range) | Median<br>G'' [kPa]<br>(range) | Median<br>G' norm<br>(range) | Median<br>G'' norm<br>(range) | Median<br>ADC<br>[10<sup>-3</sup> mm<sup>2</sup>/s]<br>(range) | Median<br>FA<br>(range) | Median<br>CBF<br>[ml/100<br>g/min]<br>(range) |
| Caudate nucleus | 1.49<br>(1.25-1.83) | 0.84<br>(0.45-1.15) | 0.82<br>(0.71-1.00) | 0.72<br>(0.40-0.95) | 0.74<br>(0.70-0.81) | 0.22<br>(0.18-0.24) | 20<br>(7-35) |
| Hippocampus | 1.83<br>(1.30-2.31) | 0.96<br>(0.72-1.14) | 1.01<br>(0.76-1.22) | 0.80<br>(0.59-0.93) | 0.88<br>(0.83-0.93) | 0.18<br>(0.16-0.23) | 20<br>(8-31) |
| Putamen | 1.78<br>(1.46-2.16) | 1.11<br>(0.77-1.58) | 0.96<br>(0.83-1.17) | 0.91<br>(0.70-1.33) | 0.73<br>(0.71-0.76) | 0.20<br>(0.17-0.23) | 24<br>(12-37) |
| Thalamus | 1.40<br>(1.21-1.55) | 0.73<br>(0.50-1.03) | 0.76<br>(0.65-0.88) | 0.60<br>(0.46-0.80) | 0.80<br>(0.76-0.84) | 0.29<br>(0.27-0.30) | 24<br>(12-44) |
| Frontal lobe | 1.65<br>(1.39-1.90) | 1.14<br>(0.88-1.33) | 0.91<br>(0.76-1.10) | 0.94<br>(0.76-1.09) | 0.88<br>(0.86-0.91) | 0.17<br>(0.15-0.18) | 44<br>(23-71) |
| Occipital lobe | 1.44<br>(1.11-2.15) | 1.17<br>(0.89-1.39) | 0.78<br>(0.61-1.20) | 1.01<br>(0.70-1.26) | 0.85<br>(0.83-0.87) | 0.16<br>(0.15-0.18) | 40<br>(25-62) |
| Parietal lobe | 1.74<br>(1.38-2.55) | 1.23<br>(0.96-1.35) | 0.95<br>(0.75-1.30) | 1.01<br>(0.81-1.16) | 0.86<br>(0.84-0.88) | 0.16<br>(0.15-0.18) | 40<br>(20-68) |
| Temporal lobe | 1.81<br>(1.55-2.28) | 1.32<br>(1.18-1.63) | 1.00<br>(0.81-1.23) | 1.10<br>(0.95-1.45) | 0.87<br>(0.85-0.89) | 0.16<br>(0.15-0.17) | 36<br>(21-60) |
| Average gray matter | 1.68<br>(1.48-1.97) | 1.16<br>(1.01-1.23) | 0.89<br>(0.81-1.07) | 0.97<br>(0.85-1.09) | 0.86<br>(0.85-0.88) | 0.17<br>(0.16-0.18) | 40<br>(22-62) |
| Average white matter | 1.84<br>(1.70-1.96) | 1.19<br>(1.10-1.32) | - | - | 0.80<br>(0.77-0.82) | 0.33<br>(0.32-0.37) | NA |

The median value of G′ in the contrast-enhancing tumor was 13 % lower than in contralateral normal-appearing white matter (cNAWM) (P<0.01). G′′ was 37 % lower (P<0.01) and FA was 52 % lower (P<0.01) in contrast-enhancing tumor than in cNAWM. ADC was 48 % higher (P<0.01) and CBF was 2.9 times higher (P<0.01) in tumor than in cNAWM. Especially CBF showed a large variability between patients, with the highest patient tumor CBF 3.2 times higher than the lowest patient tumor CBF.

In edema surrounding the tumor, median G′ was similar to cNAWM (P=0.6), while G′′ was 16 % lower than in cNAWM (P<0.01). FA was 48 % lower (P<0.05) and ADC was 53 % higher in edema than in cNAWM (P<0.01). CBF was similar in edema and cNAWM (P=0.3).

### Tumor stiffness is heterogeneous, both among patients and within tumor

The median value of G′ and G′′ was 18 % and 6 % lower in necrosis than in contrast-enhancing tumor, respectively (P<0.05). This suggests a within-tumor heterogeneity, illustrated in **Figure 2**: G′_norm_ and G′′_norm_ voxels distributions for each patient differed between contrast-enhancing tumor, necrosis and edema. The differences between patients were also larger in these regions than in normal-appearing gray matter, illustrating the interpatient tumor heterogeneity.

**Figure 2:**
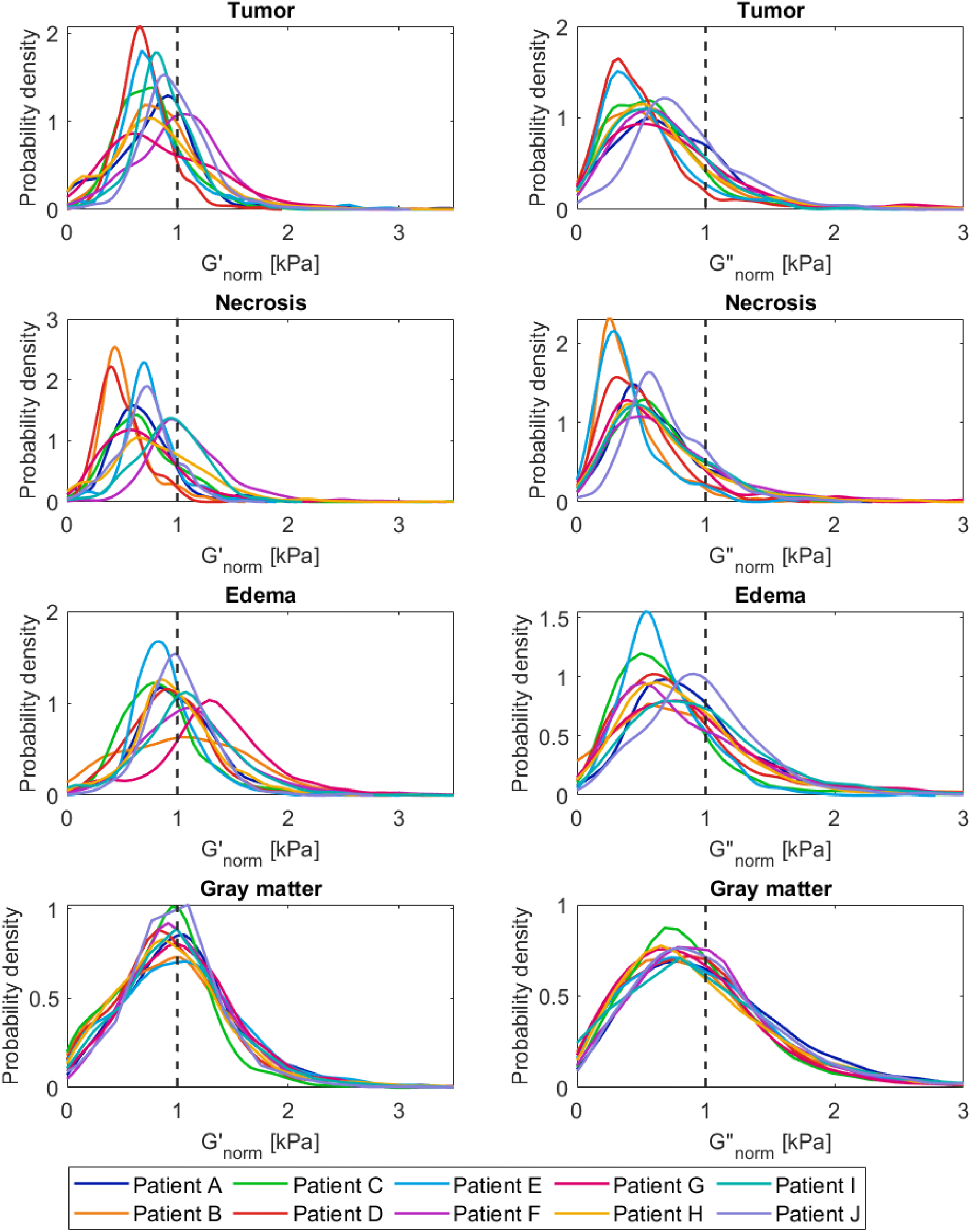
Distribution of biomechanical properties in pathological regions and gray matter. Distribution of voxel values for G′_norm_ and G′′ _norm_ in A-B) contrast-enhancing tumor, C-D) necrosis, E-F) edema and G-H) normal-appearing gray matter.

This variability is also illustrated in **Figure 3**, which shows the distribution of G′, G′′, ADC and FA among healthy subjects and patients. Measurements in tumor and edema showed a larger inter-patient variability than the values in patient white and gray matter.

**Figure 3:**
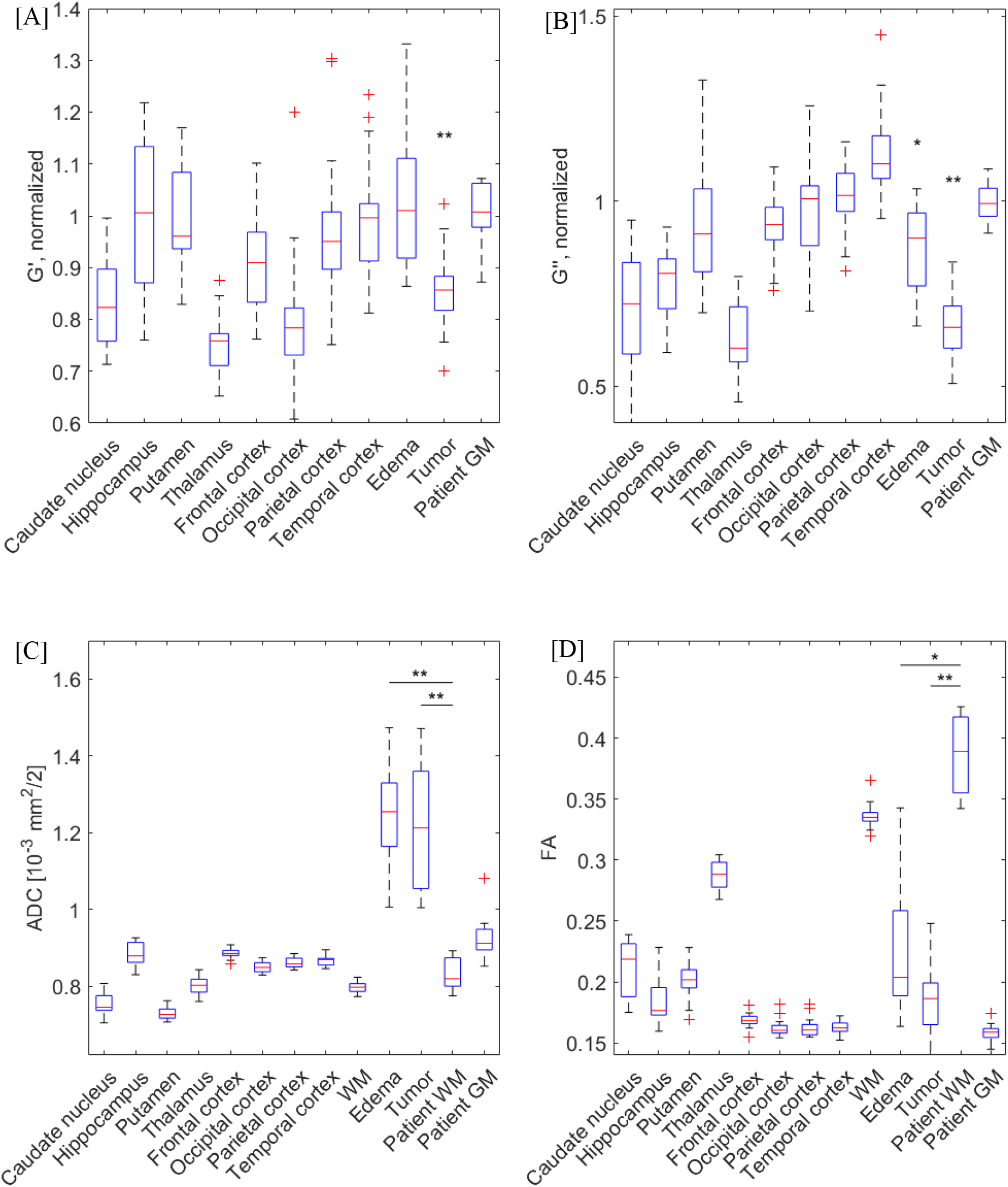
Box plots of healthy brain regions vs tumors. A) G′_norm_, B) G′′_norm_, C) ADC, and D) FA for all ROIs in healthy subjects and in edema, contrast-enhancing tumor, white and gray matter in patients. The red line shows the median value, the blue box the 25^th^ and 75^th^ percentile of the mean values, with red crosses indicating outliers. Asterisks show significant differences from patient’s cNAWM (*=P <0.05, **=P<0.01).

### Patient normal-appearing white matter shows lower stiffness, viscosity and anisotropy than white matter in healthy subjects

The white matter region in patients had 12 % lower G***′*** and G***′′*** (both P<0.001) compared to white matter in healthy subjects. White matter ADC and FA was 3 % and 16 % higher in patients than in healthy subjects (ADC: P<0.05, FA: P<0.001).

For the healthy subjects, G*′* was 9 % higher and G′′ was 39 % higher in the cortical gray matter than in deep gray matter (P<0.001). Comparing white matter to deep gray matter, G*′* was 18% higher (P<0.01) and G*′′* was 37 % higher in white matter (P<0.001). G*′* was 8% higher in white than in cortical gray matter (P<0.01), while G*′′* was similar between the two (P=0.99).

The t-SNE plot (**Fig. 4**) illustrates the separation between pathological and healthy brain regions in both patients and healthy subjects, for ADC, FA, G′_norm_ and G′′_norm_. Most regions of interest in healthy subjects tracked together, and showed a separation between deep and cortical gray matter regions. The contrast-enhancing tumor, edema and necrosis were close to each other and separated from the healthy ROIs.

**Figure 4:**
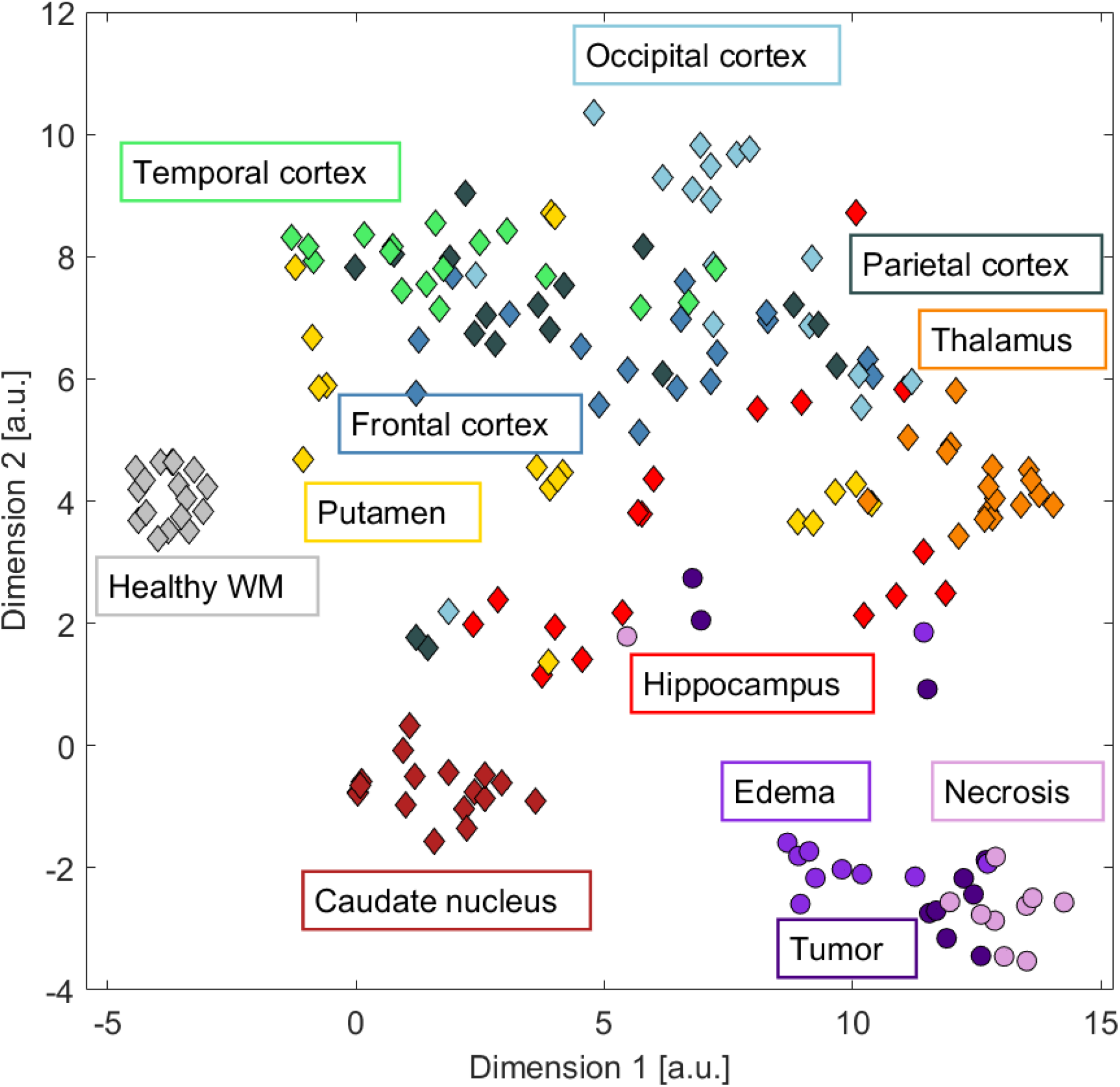
Brain regions in healthy subjects and patients with ADC, FA, G′_norm_ and G′′_norm_ reduced into two dimensions. T-distributed stochastic neighbour embedding of mean measurements, arbitrary units (a.u) on axes. Diamond markers: mean values in healthy subjects, circles: patients.

### Tissue properties approach normal values further away from tumor core

Figure 5 shows gradients of measurements moving radially out from the tumor core. For most patients, G′ and G′′ started from low values within the tumor and increased toward values found in cNAWM at the distal edge of the edema region. ADC was high in necrosis, tumor and edema, and was still 29 % (median value) higher than in cNAWM at the edema edge—although with large variation among patients. FA was low in the tumor core and increased away from it; the median value was 46 % lower at the edema edge than in cNAWM. CBF, leakage and vessel size were largest in the contrast-enhancing tumor, and gradually decreased toward the mean cNAWM value at the edema edge.

**Figure 5:**
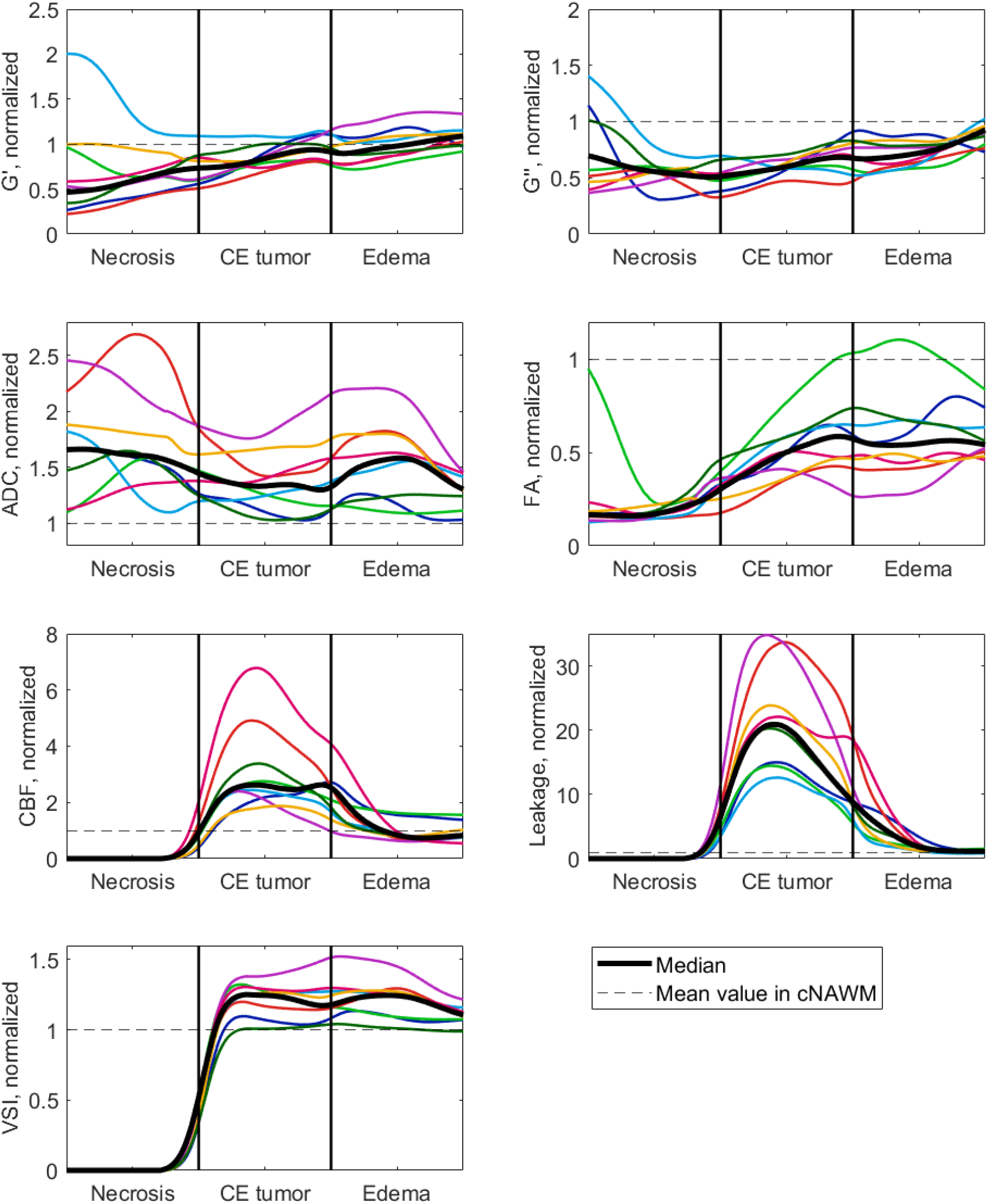
Gradients of the different measurements in the lesion, moving radially out from the tumor core. All distances were normalized to the maximum distances between necrotic core and contrast-enhancing tumor, inner contrast-enhancing tumor and edema, inner edema and edge to normal-appearing matter, respectively. Colored lines show individual patients (n=8 included), the black line shows the patient median. Abbreviation: Vessel size index (VSI).

### Tissue may have abnormal properties outside lesion area

Figure 6 illustrates gradients moving radially outward from the lesion, defined as necrosis, tumor or edema. For several of the patients, the tissue properties remained abnormal (outside the 25^th^ and 75^th^ percentile of cNAWM) for more than 5 mm into the normal-appearing tissue.

**Figure 6:**
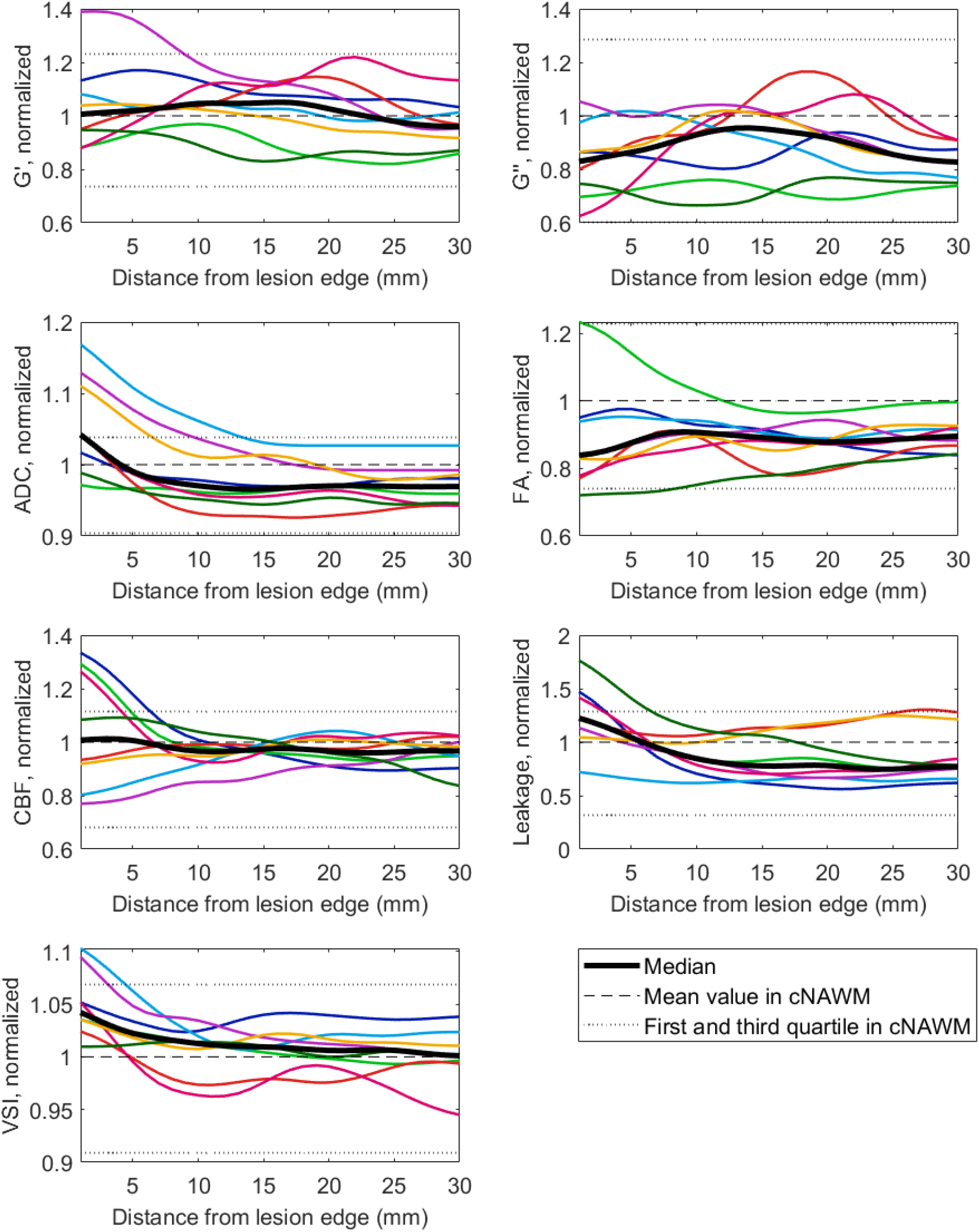
Gradients of the different measurements, moving radially outwards from the lesion edge. Measurements in tissue labelled as white matter, normalized to each patient’s cNAWM. Colored lines show individual patients (n=8 included), black line shows patient median. Abbreviation: Vessel size index (VSI).

### High cerebral blood flow related to low stiffness

A positive correlation was found between mean G*′* and G*′′* in contrast-enhancing tumor (Spearman’s rho 0.76, P<0.05) and in patient white matter (rho 0.78, P<0.05). No correlations across patients were found in the contrast-enhancing tumor between mean G*′* and mean CBF, ADC, leakage or vessel size, nor between G*′′* and the perfusion and diffusion parameters. The same was true for the other patient ROIs. For the healthy subjects, the only correlation between parameters was between G*′* and G*′′* in gray (P<0.01) and white matter (P<0.01).

We estimated voxel-by-voxel regressions to explain CBF as a function of the other measurements, all normalized to cNAWM. Our baseline model was a linear model with ADC_norm_ and FA_norm_ as predictors. Using RMSE as a criterion, we investigated how predictive power increased as we added first G′_norm_ and then both G′_norm_ and G′′_norm_ to the model. Results are presented in **Table 3**, and show that predictive power increased significantly with the inclusion of each additional predictor. After the assessment of each model, the linear model was trained using all data, and the final linear model is:

**Table 3:** Performance of models predicting CBF_norm_. Median RMSE (range) for a linear and random forest model using only diffusion parameters to predict CBF_norm_ in patients (upper row), versus including G**′**_norm_ (middle row) and both G**′**_norm_ and G**′′**_norm_ (lower row). Asterisk indicates improvement from the row above (P<0.01).

|  | Linear model | Random forest |
| --- | --- | --- |
| CBF<sub>norm</sub> as a function of ADC<sub>norm</sub>, FA<sub>norm</sub> | 0.864 (0.844-0.872) | 0.695 (0.676-0.701) |
| CBF<sub>norm</sub> as a function of ADC<sub>norm</sub>, FA<sub>norm</sub>, G' norm | 0.860 (0.840-0.869)** | 0.674 (0.654-0.680)** |
| CBF<sub>norm</sub> as a function of ADC<sub>norm</sub>, FA<sub>norm</sub>, G' norm, G'' norm | 0.859 (0.839-0.867)** | 0.626 (0.606-0.630)** |

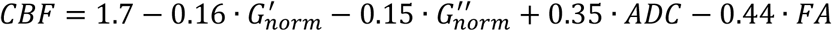

In addition to the linear model, we investigated the predictive ability of a random-forest model (**Table 3**, column 2). Again, we compared a baseline model including ADC_norm_ and FA_norm_ to models including G′_norm_, and both G′_norm_ and G′′_norm_. All three random-forest models performed better than the linear models (P<0.01). In addition, the performance of the random forest model improved by including G*′*_norm_ (P<0.01), and further improved by including both G*′*_norm_ and G*′′*_norm_ (P<0.01).

### Data accessibility

All the data and the associated meta-data generated as a part of this study is publicly available by request through Zenodo (http://doi.org/10.5281/zenodo.4926005).

## Discussion

In this study, we used MR elastography, perfusion and diffusion imaging in patients with glioblastoma and healthy subjects. We found that glioblastoma tissue was structurally degraded compared to healthy tissue in terms of all measurements. Measurements approached normal values when moving away from the tumor core, but we still found abnormal tissue properties in regions that appear normal on anatomical images. Finally, we constructed a predictive model for CBF, which showed that perfusion increased with decreased G′ and G′′.

Stiffness and viscosity in tumors, here measured by G′ and G′′, were significantly lower than in normal-appearing matter. This is consistent with previous reports: Gliomas have been found to be softer than normal tissue [4, 6, 7, 12] and substantially less viscous [4, 5, 7]. A reduction in both G′′ and G′ can be interpreted as a softening of the mechanical rigidity of the tissue [13].

Most studies of MRE in gliomas have presented mean tumor values. Streitberger et al. noted that glioblastomas were composed of stiff and soft compartments, and that the source of heterogeneity may be that glioblastomas consist of both solid masses and possibly cystic and necrotic fractions [4]. In our study, tumors were segmented into contrast-enhancing and necrotic regions. We found necrotic tumor regions to have even lower stiffness and viscosity than the contrast-enhancing parts of the tumor and the distribution of stiffness and viscosity to vary more in tumors than in normal-appearing matter. In a study of MRE in mouse-model gliomas, Schregel et al. found that tumors became softer and more heterogeneous over time with tumor progression and that softer sub-regions of the tumor were characterised by a high heterogeneity [14].

Areas displaying high FLAIR signal is presumed to represent edema due to cancer infiltration, and are considered important elements of treatment planning [9]. We found that tissue properties in edema had different characteristics than contralateral healthy tissue, which is illustrative of the tumors’ infiltrative character [15]. For some patients, abnormal tissue measurements were also found even further away from the tumor core, in regions appearing normal on anatomical scans, implying that infiltration may extend beyond the increased FLAIR signal.

The only association between the different measurements on a region level in patients was between G′ and G′′ in contrast-enhancing tumor and in white matter. The lack of correlation between biomechanical and functional properties at the region level may be due to a small sample size and low power; it may also be due to spatial variation within regions. If tissue perfusion depends on diffusion and stiffness properties, such a relationship would depend on the spatial distribution of the tumor and surrounding tissue, due to the GBM heterogeneity. Therefore, all voxels for all patients were considered when constructing regression models of perfusion as a function of the diffusion parameters. Even for a simple linear model, MRE added to the performance of the model, suggesting that MRE provides independent data. Biomechanical properties of the tissue may play an integral role in explaining the tumor vascularity. The simple linear model showed that CBF increased when G*′* and G*′′* decreased, an effect that could possibly be caused by vessels being compressed by stiff tissue and hence reduced perfusion [16]. Of course, there might be a more complex relationship between various characteristics. Other studies have found that tumor stiffness may be affected by factors such as increased cellularity, increased vessel density, and interstitial fluid pressure [6]. A preclinical study suggests that tissue stiffness is influenced by the architecture of the blood vessels, rather than their state of perfusion [17]. Further work is warranted to corroborate our findings, as understanding the mechanisms behind impaired perfusion in glioblastoma could be important for development of new treatment.

The stiffness and viscosity of the contralateral normal-appearing white matter in the patients differed significantly from the measurements in healthy subjects’ white matter. Several studies have found brain stiffness to decrease with age [18-20]. The difference between median white matter G′ between patients and healthy subjects in our study was 0.22 kPa, corresponding to -0.007 kPa/year (range 0.002-0.012 kPa/year). This amount of change is roughly comparable to earlier studies. Normalizing measurements to values in normal-appearing white matter is typically done in MRE studies of brain cancer patients [4, 5, 12]. Such a normalization can also remove confounding effects in the case of different MRE acquisition techniques [21].

Our study focused on the differences between pathological and healthy tissue, and contrast-enhancing and necrotic tumor, edema, and normal-appearing gray and white matter ROIs were used for patients. For healthy subjects, we further subdivided the gray matter regions to compare with earlier studies of MRE in the healthy brain. We found white matter to be stiffer than gray matter, consistent with earlier studies [22]. We found cortical gray matter to be stiffer than deep gray matter. This is in contrast to a large recent study, where deep gray matter was reported to be stiffer than both white and cortical gray matter [23]. A second study reported the stiffness to be lower in the deep gray matter than white matter [24], while a third study found white and gray matter stiffness to be very similar in both adults and paediatric subjects [25]. This last study also noted a data quality bias in the calculations due to the attenuation of the applied shear waves causing low MRE signal in the central regions of the brain [25, 26]. In an earlier study using this MRE method in healthy subjects, MRE data quality was found to be lower in the deep gray matter regions than in regions closer to the skull. No significant correlation between this data quality and stiffness measurements was found [11]. The existing studies differ with respect to both MRE hardware and reconstruction methods, making it challenging to conclude about the reasons for discrepancies between studies.

### Limitations

A general challenge for MR elastography is the lack of a gold standard for *in vivo* tissue stiffness measurements. A specific challenge for our study is the limited sample size, especially for patient data.

We expect some partial-volume effects with our current image resolution, possibly contributing to less precise measurements in small regions and thin structures such as the cerebral cortex. To ensure statistical validity, we only included regions with > 80 voxels. To avoid interpolation effects, MRE, diffusion and perfusion data were all analysed in their native spaces for the calculation of mean values.

Perfusion in healthy subjects was measured using ASL, as it is non-invasive, in contrast to DSC, where a gadolinium-based contrast agent is administered intravenously. This hindered direct perfusion comparison between the two groups. No EPI-distortion correction was performed for ASL images, which may have contributed to less precise coregistration to the anatomical labels.

The repeatability (coefficient of variation) of the employed MRE technique is 4 % [11]. This should be taken into account if comparing small stiffness differences between subjects.

In summary, we found that glioblastoma differed from healthy tissue in terms of G′ and G′′, CBF, ADC and FA, with heterogeneity both between patients and within tumors. Abnormal tissue properties were present in regions appearing normal on anatomical images. Finally, we showed that inclusion of MRE measurements in statistical models helped predict perfusion, with stiffer tissue associated with lower perfusion values.

## Data Availability

All the data and the associated meta-data generated as a part of this study is publicly available by request through Zenodo (10.5281/zenodo.4926005).

https://zenodo.org/record/4926005

