## Supplemental information for "Decreased tissue stiffness in glioblastoma by MR Elastography is associated with increased cerebral blood flow"

### Supplementary information

#### Image acquisition

For the patients a T1-weighted anatomical reference scan was acquired both before and after injection of a gadolinium-based contrast agent (Gadovist, Bayer Pharma AG, Germany), with a 0.1 mmol/kg body weight dose, followed by a 20 mL saline flush (BB. Melsungen AG, Germany). During contrast injection, 100 dynamic scans were acquired for dynamic susceptibility contrast (DSC) imaging [1]. Perfusion imaging in healthy subjects was performed by a 2D pseudo-Continuous Arterial Spin Labelling sequence [2].

#### Image processing

Perfusion images of the healthy subjects from ASL were analysed in the nordicICE (NordicNeuroLab AS, Bergen, Norway) software. This resulted in maps of cerebral blood flow (CBF) quantified in units of ml/100 g/min, given by the formula [2]:

$$CBF = \frac{6000 \cdot \lambda \cdot (SI_{control} - SI_{label}) \cdot e^{\frac{PLD}{T_{1,blood}}}}{2 \cdot \alpha \cdot T_{1,blood} \cdot SI_{PD} \cdot (1 - e^{-\frac{\tau}{T_{1,blood}}})} [ml/100g/min]$$

The following parameters were applied: $T_{1,blood}$= 1650 ms, labelling efficiency $\alpha$ of 0.85, and brain-blood water partition coefficient $\lambda$=0.9 ml/g [2]. Motion correction and co-registration between the ASL and the proton density images were performed prior to analysis. Because ASL in general has low sensitivity for subtle perfusion levels in white matter [2], our study includes perfusion values from deep- and cortical gray matter regions only.

The DTI and DSC images were corrected for EPI-distortion effects prior to analysis using the geometric distortion correction method FSL TOPUP [3, 4]. Thereafter, diffusion tensor imaging analysis was performed in nordicICE with motion correction, automatic detection of noise threshold and noise level cutoff. The tissue diffusivity was measured by the apparent diffusion coefficient (ADC), and the tissue anisotropy was measured by the fractional anisotropy index (FA). Patient perfusion images were also analysed in nordicICE [5] utilizing both motion- and leakage-correction on DSC data. This produced map of cerebral blood flow normalized to white matter (nCBF), leakage, and vessel size index (VSI). The two latter maps were available for eight of the ten patients.

For the MRE, phase-unwrapping and pixel-wise temporal Fourier transformation were performed on the displacement phase data in order to obtain the tissue displacement in the frequency domain. The data was filtered in image space using an 11$^{th}$ order Blackmann Harris filter [6], before the curl operator was applied in the image to eliminate information originating from the compressional wave component [7]. Through inversion, we then obtained maps of the shear storage modulus G′ and the shear loss modulus G″. To avoid artefacts from MRE reconstruction at the edge of the brain, elastography maps were eroded by 2 pixels. MRE data quality was assessed by degree of temporal nonlinearity in the MRE data, which is a measure of noise in the original phase data [8], and the ratio between the amplitude of the curl and the amplitude of the divergence [9]. This ratio quantifies the signal (curl) to noise (divergence) since the divergence of the displacement is approximately zero due to the incompressible nature of tissue. All scans had a mean curl-divergence ratio above 5 and any voxels with nonlinearity above 50 % were excluded in the MRE maps.

#### Image registration and analysis

*Segmentations of patient data*

Segmentations of contrast-enhancing tumor, edema, necrosis, and normal-appearing gray and white matter was performed automatically using a convolutional neural network (CNN). This CNN is based on the 3D-Unet architecture defined by Juan-Albarracín et al. [10], trained with 262 BRATS [11, 12] pre-surgical exams and 222 follow-up exams from our institution. The segmentations were done for each patient based on pre- and post-contrast T1-weighted images, T2-weighted and FLAIR images. Contrast-enhancing and necrotic tumor were defined as the enhancing and non-enhancing tumor region on post-contrast $T1_w$ images, respectively. Edema was defined as the hyperintense region on the FLAIR images. Gray- and white-matter masks were eroded by one voxel, and only the opposite hemisphere of the brain from the tumor was used for the normal-appearing gray- and white-matter masks. For the cases where the tumor affected both hemispheres, a 3 cm margin from the distal edge of the tumor and edema was used for the gray and white matter masks.

For comparisons between patients on an image voxel level, and for calculations of gradients outward from the patients, all maps were registered to the Montreal Neurological Institute (MNI) space [13], by means of an affine transformation and using a nearest-neighbour interpolation to preserve image voxel integrity. For the regression analysis on the voxel level, all maps were smoothed with a 3D Gaussian filter with sigma 1. The analysis was performed in Matlab (version R2021a, MathWorks, Natick, MA, USA).

To assess the spatial distribution of the parameters in necrosis, the distance to contrast-enhancing tumor was calculated for all points in the necrosis, and normalized to the maximum distance to contrast-enhancing tumor. This was repeated in contrast-enhancing tumor, using the distance to edema, and in edema, using the distance to normal-appearing tissue. For visualization, the gradients were smoothed using a Gaussian-weighted moving average filter of window length 70.

In order to make gradients outward from the lesion edge, here defined as the distal edema, an ROI consisting of necrosis, contrast-enhancing tumor and edema was dilated 1 mm at a time, 30 times. For each dilation, the last ROI was subtracted and the mean value of each new layer calculated. The gradients were smoothed using a Gaussian-weighted moving average filter of window length 10.

*Regions of interest in healthy subjects*

All image registrations in healthy subjects were performed using Matlab and SPM12 (version 7487, Wellcome Trust Centre for Neuroimaging, London). First, anatomical T1-weighted images were warped to the MNI brain region template [14], and the inverse deformation fields were used to reorient the binary maps of the brain regions of interest into the each subject’s anatomical space. T1-weighted images and labels were then co-registered by affine transformations, using nearest-neighbour interpolation, into the native spaces of the MRE, DTI and the ASL images.

ROIs in the healthy subjects were extracted from the MNI template. The deep gray matter regions included in our study were head of the caudate nucleus, putamen, thalamus and hippocampus. The cortical gray matter regions included were the frontal lobe, the occipital lobe, the parietal lobe and the temporal lobe. For MRE and DTI images, a white matter ROI was also included. ADC maps from the diffusion acquisition was used to reduce any potential errors from partial volume effects. A mask with a cut-off ADC value of < 1.2 $\times 10^{-3}$ $mm^2/s$ was used to exclude voxels with free diffusion from DTI and MRE maps, to ensure no voxels with cerebrospinal fluid were included [15].

[11] B.H. Menze, A. Jakab, S. Bauer, J. Kalpathy-Cramer, K. Farahani, J. Kirby, Y. Burren, N. Porz, J. Slotboom, R. Wiest, L. Lanczi, E. Gerstner, M.A. Weber, T. Arbel, B.B. Avants, N. Ayache, P. Buendia, D.L. Collins, N. Cordier, J.J. Corso, A. Criminisi, T. Das, H. Delingette, C. Demiralp, C.R. Durst, M. Dojat, S. Doyle, J. Festa, F. Forbes, E. Geremia, B. Glocker, P. Golland, X. Guo, A. Hamamci, K.M. Iftekharuddin, R. Jena, N.M. John, E. Konukoglu, D. Lashkari, J.A. Mariz, R. Meier, S. Pereira, D. Precup, S.J. Price, T.R. Raviv,

S.M. Reza, M. Ryan, D. Sarikaya, L. Schwartz, H.C. Shin, J. Shotton, C.A. Silva, N. Sousa, N.K. Subbanna, G. Szekely, T.J. Taylor, O.M. Thomas, N.J. Tustison, G. Unal, F. Vasseur, M. Wintermark, D.H. Ye, L. Zhao, B. Zhao, D. Zikic, M. Prastawa, M. Reyes, K. Van Leemput, The Multimodal Brain Tumor Image Segmentation Benchmark (BRATS), IEEE Trans. Med. Imaging 34(10) (2015) 1993-2024.
[12] S. Bakas, H. Akbari, A. Sotiras, M. Bilello, M. Rozycki, J.S. Kirby, J.B. Freymann, K. Farahani, C. Davatzikos, Advancing The Cancer Genome Atlas glioma MRI collections with expert segmentation labels and radiomic features, Sci Data 4 (2017) 170117.
[13] M. Brett, I.S. Johnsrude, A.M. Owen, The problem of functional localization in the human brain, Nat. Rev. Neurosci. 3(3) (2002) 243-9.
[14] N. Tzourio-Mazoyer, B. Landeau, D. Papathanassiou, F. Crivello, O. Etard, N. Delcroix, B. Mazoyer, M. Joliot, Automated anatomical labeling of activations in SPM using a macroscopic anatomical parcellation of the MNI MRI single-subject brain, Neuroimage 15(1) (2002) 273-289.
[15] J.L. Bykowski, L.L. Latour, S. Warach, More accurate identification of reversible ischemic injury in human stroke by cerebrospinal fluid suppressed diffusion-weighted imaging, Stroke 35(5) (2004) 1100-6.
